# A Public Health Hackathon for Medical Students in Africa: Process, Outcome and Recommendations

**DOI:** 10.1101/2023.01.28.23284802

**Authors:** Abdulhammed Opeyemi Babatunde, Farida Oluwabukola Brimmo, Umulkhairah Onyioiza Arama, Madu Gloria Onyinyechi, Kasule Aiden Josephat, Austine Oluwatobi Osiene

## Abstract

Africa has a dual burden of disease, which causes preventable morbidities and fatalities. This is a result of our healthcare system’s deficiencies, which has suffered a serious decline since the COVID-19 pandemic. However, this opened up the possibilities for digital health interventions, which innovators could utilize to provide solutions to these public health issues. Health hackathons, which offer an environment for innovators to brainstorm and collaborate, are rare in Africa. This paper outlines the planning and execution of a virtual hackathon and explores its implications for the promotion of public health in Africa.

Over the course of a month, we collaborated with innovation hubs in Africa to hold an open call, a training session, a design sprint, as well as a 48-hour virtual hackathon. We received 68 submissions from 13 African nations. Following the selection of 10 teams, design thinking was employed to develop solutions to a public health problem in Africa. The theme for the hackathon was ‘Promoting Health Equity with Digital Technology in Africa’ and areas of focus were non-communicable diseases; infectious disease epidemiology and surveillance; and health information and data management. All ten teams developed prototypes, the top three teams were offered the opportunity to continue on to the start-up accelerator program, while the winning team also received a cash prize.

In conclusion, the public health hackathon challenged African medical students to develop innovations for healthcare problems. There is a need for further study to evaluate the solutions developed.

## BACKGROUND

Africa faces a range of public health challenges resulting in preventable mortalities and morbidities from both communicable and non-communicable diseases (Feldscher, 2020). High disease burden and preventable deaths in Africa threaten the feasibility of achieving Africa’s developmental blueprint by 2063 (Nkengasong & Tessema, 2020). The continent accounts for 94%, 73%, and 25% of the global burden of malaria, HIV/AIDS, and Tuberculosis respectively (WHO, 2020). In addition, there is a growing burden of non-communicable diseases such as cancer, cardiovascular diseases, chronic kidney disease, mental and substance use disorders, and diabetes among the African population (WHO Africa, 2022). The rate of mortality from NCDs is predicted to be highest in sub-Saharan Africa due to late detection, poor management, lack of health insurance, and weak healthcare system (Dalala et al, 2011; Checkley et al, 2014).

The COVID-19 pandemic has also further worsened the healthcare system in African countries (WHO Africa, 2020). However, it creates opportunities for healthcare developments through research and innovations, especially digital health. Digital health interventions have shown to help reduce disease burden, facilitate access to care and reduce health inequalities (Boutilier et al, 2022).

Hackathons provide an opportunity for innovators to brainstorm creative ideas to solve a defined problem. Historically, hackathons mainly targeted the tech industries (Poncette et al, 2020). However, in recent times, they have been conducted to develop sustainable health innovations by fostering interdisciplinary and user-centred approaches. Health hackathons are still very uncommon in African countries compared to developed countries despite facing disproportionately greater health challenges (Bhandari et al, 2017). Hence, the Federation of African Medical Students’ Associations (FAMSA) Standing Committee on Medical Education and Research (SCOMER) organized her maiden edition of Public Health Hackathon for African medical students to solve public health problems facing African communities. The theme for the hackathon was ‘promoting equity with digital health technology in Africa’ with four focus areas which include; Infectious Disease Epidemiology and Surveillance; Non-Communicable Diseases; Health Information and Data Management; and Genomics and Precision Medicine. The objectives of the hackathon include (i) to develop teamwork, critical thinking, and problem-solving skills of African medical students (ii) to challenge African medical students to present creative and innovative solutions to problems of Africa’s healthcare system (iii) to contribute towards the improvement of health and wellbeing of African communities.

This paper aims to describe the planning and implementation of the virtual hackathon, summarise the participating teams, and discuss implications for public health promotion in Africa.

## METHODS

### Methods

We combined an open call, training and design sprint. The hackathon committee comprised of eight FAMSA SCOMER officers representing different medical schools in Africa. The committee was involved in the overall planning and execution of the project including sourcing for partners, mentors, reviewers, sponsors, and trainers. The 48 hours hackathon held on October 7-8, 2022 and final pitching ceremony on October 9, 2022. We used used a variety of technologies for internal and external interactions, including Zoom, Gmail, WhatsApp, and Google Meet. Each team was expected to improve on the submitted idea using design thinking process to develop a prototype and create a 5-minute pitch for presentation to panel of judges. Design thinking is a problem-solving approach which is human-centered which involve empathizing, defining, ideating, prototyping and testing in order to develop or improve a product that addresses the needs of the end-user (Gottgens & Oertelt-Prigione, 2021). The final pitching event took place on October 9, 2022, and each team pitched their innovation to a panel of judges via the Zoom platform.

### Hackathon resources

The first and most important resource was the partnership with Co-creation Hub (CcHub), an organisation that provides an environment for social entrepreneurs, tech companies, investors and hackers to co-create innovative solutions to societal problems. CcHub has vast experience in co-creating health tech innovations in Nigeria and other African countries. Hence, their contribution was pivotal at every stage of the planning and during the hackathon. Other partners and helpful resources include One Health Tech, The Unicorn Den, The Innovation and Design thinking Academy (TIDA) and 4 Youth By Youth.

### Phase I: Open Call

The theme of the open call was the same as that of the hackathon. Applicants were permitted to apply as an individual or in teams of 3–5 members, with 75% being medical students from any university in Africa. The open call was publicized on all FAMSA social media, in newsletters, and in WhatsApp groups through the liaison officers. We organized a webinar to explain the scope of the hackathon and provide step by step guide on the open call submission. The recording of the webinar was disseminated across all social media platforms. To register, applicants were asked to fill out an online Google form which included personal information, skillset, project name, problem, idea, and team. Individuals without any project ideas but with skill sets were also allowed to apply but were to be paired with others. The call lasted for a month with a three-day extension for late submissions.

### Phase II: Hackathon

The submitted entries were internally reviewed by two members of the organising committee to confirm eligibility and remove all identifiers before sharing equally among five external reviewers. The submissions were reviewed and graded based on the criteria: clarity, relevance, novelty and creativity, feasibility and sustainability, promotion of equity, and digital technology **(Supplementary Material 1)**. The top 7 ‘team’ entries and 3 ‘individual’ entries were selected and invited to participate in the 48 hours hackathon. The selected individual entries were paired with four other applicants with complementary skill sets to form a team each. Ten teams of 3-5 members each were invited to the hackathon and received one-on-one mentorship from health innovators from different African countries. Each team was expected to have at least 2 meetings with their mentor. On the first day, we held an introductory session to welcome the teams, introduce them to their mentors, and go over the hackathon’s guidelines and expected deliverables. This was followed later in the day by the first workshop sessions, which were facilitated by Co-creation hub. The sessions’ titles were “Innovation in Healthcare” and “Ideation, Prototyping, and Pitching.” Meetings with team mentors and hacking sessions took up the rest of the day. The second workshop session which was held on the morning of day two was facilitated by The Innovation and Design thinking Academy. Teams received training on business model canvas and user-journey map design. At the end of the second day, each team presented a demo pitch to the organizing committee. The final pitching ceremony was held on 9th October 2022 on Zoom. Participants had five minutes to present their ideas to three independent expert judges, with an additional five minutes allotted for questions and answers. The panel of judges consisted of a design thinking and business consultant; a public health expert with experience in managing health hackathons and open call; and a medical doctor and health innovator. The teams were accessed using a pre-set judging criteria which was developed by the organising committee and validated by the judges. **(Supplementary Material 2)**:

Following the announcement of results, all teams were acknowledged and awarded certificates of participation. The top three teams were given the opportunity to proceed to The Unicorn Den, a startup accelerator program. Additionally, the winning team was awarded a cash prize of USD 200 for prototype development.

**Figure 1:**
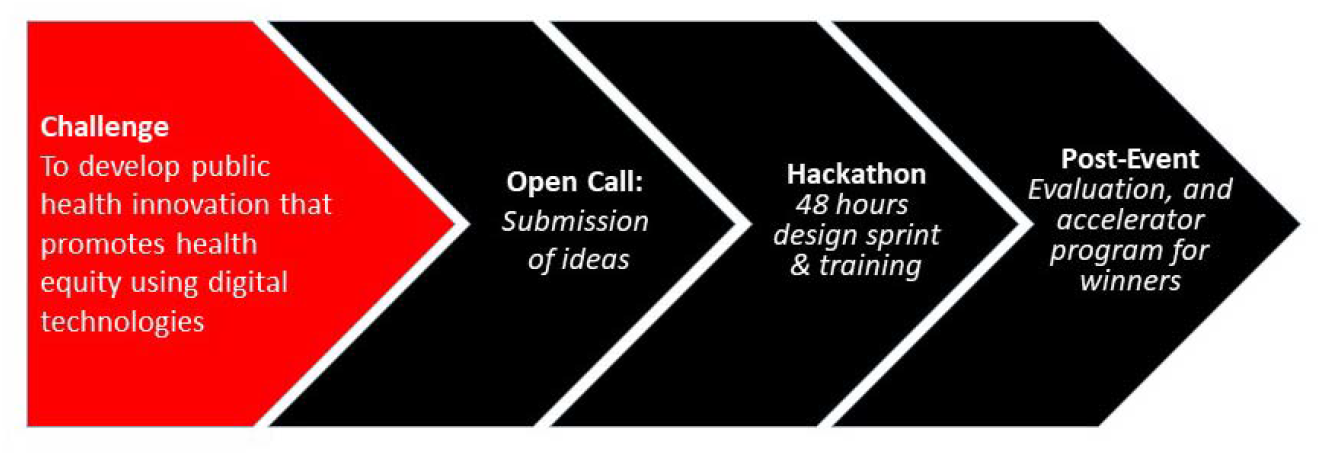
Stages of the Public Health Hackathon.

## RESULTS

In total, we received 68 submissions (26 teams and 42 individuals) from the open call from 13 African countries (Nigeria 49; Zimbabwe 1; Ghana 1, Cameroon 2, Somalia 1, Tanzania 4, Zambia 1, South Africa 1, Liberia 1, Uganda 1, Kenya 3, Mali 1, Sudan 2). 59 (86.8%) entries were submitted by medical students. Table 1 summarizes the ten teams that participated in the 48 hours hackathon.

**Table 1:** Summary of the ten teams at the hackathon.

| Team ID | Members | Countries | SubTheme | Idea | Score |
| --- | --- | --- | --- | --- | --- |
| Team 1 | 3 | Nigeria | Non- | Increasing | 92.0% |
|  |  |  | Communicable Diseases | access to blood supply |  |
| Team 2 | 5 | Nigeria, Cameroon, Tanzania | Health Information and Data Management | Online platform for chronic disease patients to connect and share information | 81.5% |
| Team 3 | 5 | Nigeria | Infectious Disease Epidemiology and Surveillance | Improve case reporting and disease surveillance in rural areas | 80% |
| Team 4 | 4 | Nigeria | Health Information and Data Management | Improve collection and storage of medical records | 79.5% |
| Team 5 | 4 | Nigeria | Health Information and Data Management | Improve handling and transfer of medical record electronically | 77.5% |
| Team 6 | 5 | Nigeria, Somalia, Zimbabwe, Ghana | Infectious Disease Epidemiology and | Using storytelling for public health | 74.5% |
|  |  |  | Surveillance | education about infectious diseases |  |
| Team 7 | 5 | Nigeria | Non-Communicable Disease | Improve monitoring and management of hypertension | 74.0% |
| Team 8 | 5 | Kenya | Health Information and Data Management | Increase access to emergency care with GPS | 71.0% |
| Team 9 | 5 | Nigeria, Kenya, Zambia, Sudan | Health Information and Data Management | A messaging app for smooth doctor-patient interaction | 70.5% |
| Team 10 | 3 | Tanzania | Health Information and Data Management | Provide decentralized, patient-owned electronic medical record | 69% |

All the innovations involved the use of mobile health technology (mHealth). Most (6 out of 10) of the hackathon ideas focused on health information and management. The average score of the teams after the pitch ranged from 69% to 92%. The winning team, Team 1, developed a prototype for a mobile application and web application that addresses the shortage of blood in Africa. The digital technology connects patients, blood donors, blood banks, and hospitals across Africa using artificial intelligence and blockchain to ease the process of accessing compatible blood types in real-time. The innovation is being piloted in Nigeria before scaling across Africa. The second-place team, Team 2, developed a prototype of a mobile application and website to connect people living with chronic diseases to anonymously discuss extensively about their health conditions, seek and receive advice from existing patients, and even specialist healthcare generating data in the process for research and decision-making. The third-place team, Team 3, developed a prototype for a web app that allows easy reporting of emerging diseases in rural communities directly into the national surveillance system with a solar-powered tablet. All other participating teams pitched ideas that employ digital health technology to promote public health in different African communities.

Following the hackathon, an online anonymous evaluation form was shared with all participants, mentors, judges, and reviewers for comment on the hackathon and recommendations for future hackathons. Only 13 respondents (all participants) were received from the survey. All respondents gave positive remarks. One of the participant’s remarks is below;

“*It was an experience to remember. I loved the way the groups were created and how we had to work in teams towards the hackathon finale. There was so much I learned during the final day, even though it was my first time. I believe the connections we made during this hackathon will be vital*.”

## DISCUSSION

We organized the first public health hackathon involving medical students in Africa using crowdsourcing. The hackathon provided an opportunity for future healthcare professionals to collaborate and brainstorm innovative solutions to public health challenges in Africa with training and mentorships from health innovators across Africa. The identified challenges were inspired by participants’ understanding of local health care and interactions with affected populations. The hackathon resulted in ten public health innovations and prototypes (mostly low-fidelity) that address public health issues and are ready for pilot testing in various African countries. The innovations were judged by an independent panel based on clarity, relevance, novelty, feasibility, promotion of equity and team. It exposed participants to the concept of human-centred design thinking for developing sustainable innovation in healthcare. The hackathon also prioritised the promotion of health equity by using digital health technology to provide access to health information and services for everyone. It was also an opportunity to connect medical students from different African countries to industry experts and mentors for a lasting relationship.

Students at all levels of education should be instructed and encouraged to use creative and entrepreneurial techniques when dealing with the complex difficulties of today’s health system in order to generate the next generation of healthcare innovators. Using innovation and entrepreneurship training to handle complicated challenges and emphasis on solution creation is becoming more widespread in medical campuses throughout the world. Hackathons in health education systems can promote and enhance active learning, creative thinking, interdisciplinary teamwork, innovation and the generation of fresh ideas. Numerous knowledge processes are generated for sharing, synthesizing, and inventing. It creates a learning envronment that allows students to use factual and procedural knowledge in humorous, stimulating, and collaborative ways. In addition, hackathons allow students to practice pitching presentation skills that are now essential in all areas of professional life.

### Limitations

The hackathon had some challenges and limitations. First, it was entirely virtual which denied some potential participants without an internet-enabled device the opportunity to participate. Although the target population was medical students who are mostly digitally savvy and have access to digital devices. However, internet connectivity, power supply, and their associated costs were major challenges that hindered the active participation of some team members and mentors. It also interfered with the judging, as judges might miss out on some information due to unstable internet connectivity on both ends. The pitch deck was shared with the panel of judges about 24 hours before the pitch to provide a background understanding of each team’s idea. Besides, the duration of the hackathon was too short to develop and validate a comprehensive solution that is user-centred and feasible in the target population. Also, the judging criteria, such as feasibility were not supported with outcome data but were mainly based on the discretion of the judges. Due to lack of funding, the winning teams were not supported with seed-funding to drive their innovations thereby making implementation challenging.

## RECOMMENDATION FOR FUTURE HACKATHON

The public health hackathon is a novel initiative by the Federation of African Medical Students’ Associations to challenge medical students to rise up and solve problems facing the healthcare system in Africa. It allowed young people to lead the development of innovations that would disrupt the healthcare sector by using digital health technology to promote health equity. We recommend that future hackathons should be physical; however, due to lack of resources and logistics, the virtual hackathon should provide internet support for participants, mentors, and judges. Also, the organizing committee should invite investors, philanthropists, global health leaders and international organisations to provide seed-funding for winning teams for implementation. Lastly, future hackathons should allow equal participation of students and professionals from other disciplines to foster interdisciplinary collaboration. The period of the hackathon might also be longer to provide an opportunity for adequate market research, user feedback, and prototype testing. The process of planning and results from this hackathon can serve as a template for future hackathons by FAMSA and other organisers of health hackathons in Africa. There is a need for further study to evaluate the effectiveness of this approach and formulate a guide for health hackathons in Africa.

## CONCLUSION

We described the process and outcomes of the first medical students’ public health hackathon in Africa that focused on building innovative solutions to promote health equity using digital health technologies in African communities. The hackathons provide opportunities for young people and future healthcare professionals to develop sustainable health innovations capable of reducing the disease burden in Africa.

## Data Availability

All data produced in the present study are available upon reasonable request to the authors

## ACKNOWLEDGMENTS

We appreciate the dedication of the hackathon participants to their projects, the enthusiasm and commitment of the mentors and judges to innovation and design thinking, and the support of our partners in making the hackathon a success.

## CONTRIBUTORS

All authors contributed to the delivery of this project. AOB wrote the first draft of the manuscript, which was later edited by all authors.

**Supplementary Material 1: Judging criteria for open call**

**Supplementary Material 2: Judging criteria for hackathon**

## REFERENCES

Bhandari A, Hayward M, Luminary Labs MIT Hacking Medicine. [2017-06-08]. Health hackathon database http://hackingmedicine.mit.edu/health-hackathon-database/ [Ref list]

Boutilier, J. J., Yoeli, E., Rathauser, J., Owiti, P., Subbaraman, R., & Jónasson, J. O. (2022). Can digital adherence technologies reduce inequity in tuberculosis treatment success? Evidence from a randomised controlled trial. BMJ global health, 7(12), e010512. https://doi.org/10.1136/bmjgh-2022-010512

Checkley W, Ghannem H, Irazola V, et al. Management of NCD in low- and middle-income countries. Glob Heart. 2014;9(4):431–443. doi:10.1016/j.gheart.2014.11.003

Dalal S, Beunza J, Volmink J et al. Non-communicable diseases in sub-Saharan Africa: what we know now. Int J Epidemiol. 2011; 40: 885–901

Feldscher K. Africa CDC head is driving a new public health agenda on the continent. https://www.hsph.harvard.edu/news/features/africa-cdc-head-is-driving-a-new-public-health-agenda-on-the-continent/. March 12, 2020 (Accessed 14th June, 2022)

Göttgens I, Oertelt-Prigione S. The application of human-centered design approaches in health research and innovation: a narrative review of current practices. JMIR Mhealth Uhealth. (2021) 9(12):e28102. 10.2196/28102.

Nkengasong JN, Tessema SK. Africa Needs a New Public Health Order to Tackle Infectious Disease Threats. Cell. 2020;183(2):296–300. doi:10.1016/j.cell.2020.09.041

Poncette AS, Rojas PD, Hofferbert J, Valera Sosa A, Balzer F, Braune K. Hackathons as Stepping Stones in Health Care Innovation: Case Study With Systematic Recommendations. J Med Internet Res. 2020;22(3):e17004. Published 2020 Mar 24. doi:10.2196/17004

WHO Africa. Deaths from noncommunicable diseases on the rise in Africa. 11 April 2022. https://www.afro.who.int/news/deaths-noncommunicable-diseases-rise-africa

WHO Africa. Covid-19 hits life-saving health services in Africa. 5th November, 2020. https://www.afro.who.int/news/covid-19-hits-life-saving-health-services-africa

WHO. Global tuberculosis report 2020. Geneva: World Health Organization, 2020.

