## Supplementary material 1 for "A Public Health Hackathon for Medical Students in Africa: Process, Outcome and Recommendations"

**FAMSA SCOMER’S PUBLIC HEALTH HACKATHON**

**JUDGE’S SCORE SHEET: ROUND 1**

**TEAM NAME:**

**ENTRY NUMBER:**

**JUDGE'S INSTRUCTIONS:**

*For each bullet point, use the SCORING LEGEND to award points corresponding to your evaluation of the student's or team's written work. You should award points based on the questions for each section and the following overall criteria: (1) Clarity of the idea, (2) Relevance of the solution and (3) Novelty and creativity (4) Feasibility and Sustainability, (5) Promotion of equity and (6) Digital Technology*

*For each section, 1-6, add the points for all bullets and put total in the "Section Total" box.*

**CLARITY OF THE IDEA**: The idea is clear, and presents a compelling answer to the question “What value will you create and for whom?”

| How well does the written work address the following? | **Points possible** | **Points awarded** | **SECTION TOTAL** |
| --- | --- | --- | --- |
| - The student/ team has clearly described their product/service idea | **4** |  |  |
| - The student/ team has clearly identified their target market | **2** |  |  |
| - The student/team has given evidence that users in the target market demand or desire their product/service | **2** |  |  |
| - The student/ team has clearly stated the benefits their product/service offers the target market and how those benefits are offered | **2** |  |  |

**Comments:**

**RELEVANCE OF THE SOLUTION**: The solution actually addresses a very important problem in the African community

| How well does the written work address the following? | **Points possible** | **Points awarded** | **SECTION TOTAL** |
| --- | --- | --- | --- |
| - The student/ team has identified a very significant “need” or “want” | 5 |  |  |
| - The student/team has identified and demonstrated in depth understanding of a key problem in the African health sector | 5 |  |  |
| - The student/team product/service idea has adequately addressed the key problem for the target population | 5 |  |  |
| - The student/team has presented a realistic strategy for their solution to lead to impact | 5 |  |  |

**Comments:**

**NOVELTY AND CREATIVITY**: The solution is creative and unique either to the context in which it is implemented or the geographical location

| How well does the written work address the following? | **Points possible** | **Points awarded** | **SECTION TOTAL** |
| --- | --- | --- | --- |
| - The student/ team identifies existing products/ services | **5** |  |  |
| - The student/team explained how their product/service idea is an improvement from existing or soon to be available products/ services | **10** |  |  |
| - The student/team cited evidence to demonstrate their competitive advantage | **5** |  |  |

**Comments:**

**FEASIBILITY AND SUSTAINABILITY**: The solution is realistic and practical and impacts stated are based on the capacity of the team, resources and technicality.

| How well does the written work address the following? | **Points possible** | **Points awarded** | **SECTION TOTAL** |
| --- | --- | --- | --- |
| - The solution is financially self-sustainable within a short while after launch | **10** |  |  |
| - The solution address operational, interventional and identity sustainability | **10** |  |  |
| - The technicality, utility and business model of solution fit the target population | **10** |  |  |

**Comments:**

**PROMOTION OF EQUITY**: The solution breaks the barrier of health inequality and provide access to everyone including remote communities.

| How well does the written work address the following? | **Points possible** | **Points awarded** | **SECTION TOTAL** |
| --- | --- | --- | --- |
| - Solution promote health of vulnerable groups e.g children and women, refugee, less privilege, etc | **4** |  |  |
| - Reduce cost of healthcare or financial burden of healthcare | **2** |  |  |
| - Improve access to healthcare for remote communites and resource limited settings | **2** |  |  |
| - Improve quality of healthcare | **2** |  |  |

**Comments:**

**DIGITAL TECHNOLOGY:** The solution leverages on technologies to improve healthcare quality and access.

| How well does the written work address the following? | **Points possible** | **Points awarded** | **SECTION TOTAL** |
| --- | --- | --- | --- |
| - Solution promote digital health e.g SMS, mobile app, website etc | **4** |  |  |
| - Choice of digital technology suit target population | **3** |  |  |
| - Solution consider populations without internet or mobile phones | **3** |  |  |

**Comments:**

**Final score for written document: _________/100 Points (Maximum)**
