## Supplementary material 2 for "A Public Health Hackathon for Medical Students in Africa: Process, Outcome and Recommendations"

**Pitch Evaluation Form final**

**Team Name:**

**Judge Name:**

**Clarity (15%)**

**Is the idea clear, and presents a compelling answer to the question “What value will you create and for whom?**

1.

**Relevance (15%)**

**Does the team understand the problem and does the solution actually addresses a very important health problem in the selected African community?**

1. .

**Novelty (15%)**

**The solution is creative and unique either to the context in which it is implemented or the geographical location? Does the team understand their competitors and competitive advantage?**

**Feasibility, Scalability/Replicability or Sustainability (25%)**

**Is the solution realistic and practical and impacts stated are based on the capacity of the team, resources (including identified partners) and technicality? Is it sustainable, replicable elsewhere and scalable? Is the team already implementing?**

1.

**Promotion of Equity and Fairness with digital tech (15%)**

**Does the solution breaks the barrier of health inequality and provide access to everyone including remote communities with digital technology? Does the team include vulnerable groups and paid attention to people without internet or mobile phones also?.**

1.

**Team (15%)**

**Is the team diverse in term of skills, capacity to implement their idea, and gender equality**

1.

**Total:**

Comment:
